# A framework for estimating cardiorespiratory fitness from diverse exercise tests

**DOI:** 10.1101/2025.11.05.25339523

**Authors:** Tomas I. Gonzales, Nick Wareham, Soren Brage

## Abstract

Comparisons of cardiorespiratory fitness across population studies are hindered by a lack of standardisation in exercise testing and reliance on estimation methods that assume steady-state exercise. We developed a generalised framework that standardises maximal oxygen consumption 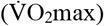 estimation by harmonising dynamic heart rate modelling with individualised calibration of exercise energetics. We constructed and evaluated the framework using submaximal and maximal tests (treadmill walking and running, cycle ergometer, stepping, overground walking) from 911 adults. Agreement between framework estimates and directly measured 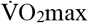 was strong (non-significant bias, Pearson’s r=0.80). We then applied both framework and canonical methods to estimate 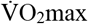 in 11,307 UK Fenland Study adults and examined associations with cardiometabolic risk. Framework-estimated 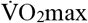 was more strongly associated with blood glucose, fasting insulin, and high-density lipoprotein cholesterol (combined risk factor model AIC=125,743) compared to canonical estimates (AIC=129,227). The framework standardises fitness estimation across diverse tests, yielding a more reliable and comparable biomarker of cardiometabolic health.

## INTRODUCTION

Accurate assessment of cardiorespiratory fitness is crucial for understanding its relationship with health outcomes in population research ^1^. However, variability in exercise testing – such as the use of non-steady-state tests and different modalities (e.g., treadmills in some studies, cycle ergometers in others) – and limitations of traditional 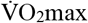 estimation methods hinder the comparability of findings from subpopulations within the same study ^2^ or across different studies ^3–5^. This is because traditional methods often rely on assumptions of steady-state exercise and may not fully capture individual physiological variability in heart rate response, metabolic efficiency, and recovery kinetics. These limitations not only weaken the conclusions drawn from large-scale population studies but also constrain our ability to develop individualised public health interventions.

To address this challenge, we present a generalised modeling framework that standardises cardiorespiratory fitness estimation for exercise testing. The framework harmonises foundational principles from exercise physiology ^6^ with dynamic modeling of exercise heart rate response ^7–11^ to create a model that accounts for non-steady-state exercise and individual physiological variability. This approach allows for the estimation of 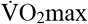 by individually calibrating the energetic demands of different exercise modalities. By leveraging these features, the framework could advance health research by providing a standardised approach to cardiorespiratory fitness assessment that would enable robust comparisons of fitness across populations with diverse exercise test data.

To validate the framework and demonstrate its utility, we pursued a multi-stage approach. First, we constructed and tuned the framework using a pooled dataset encompassing a wide variety of exercise tests and protocols. Next, we established its criterion validity on a hold-out sample by examining the accuracy of its 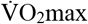 estimates against directly measured values. We further interrogated its physiological plausibility by comparing other parameter estimates to values reported in the literature. Finally, to establish its construct validity, we applied the framework to the UK-based Fenland study ^12,13^ and compared associations with cardiometabolic risk factors against those obtained from two traditional estimation methods.

## RESULTS

### Study overview and data sources

**Figure 1.** provides a flow diagram of the study design. To develop the fitness estimation framework, we pooled exercise test data from three sources: the Biobank validation study (Cambridge, UK), the Step test validation study (Cambridge, UK), and a repository of exercise test data on PhysioNet (**Table 1**). The pooled dataset, encompassing a diverse range of exercise modalities and protocols from both submaximal and maximal tests with directly measured 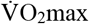, was split into derivation (80%), validation (10%), and test (10%) subsets. We used the derivation subset to develop the framework, the validation subset to tune its hyperparameters, and the test subset to evaluate its performance. We then applied the framework to exercise test data from the UK-based Fenland cohort study to examine associations between estimated 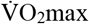 and cardiometabolic risk factors.

**Figure 1.**
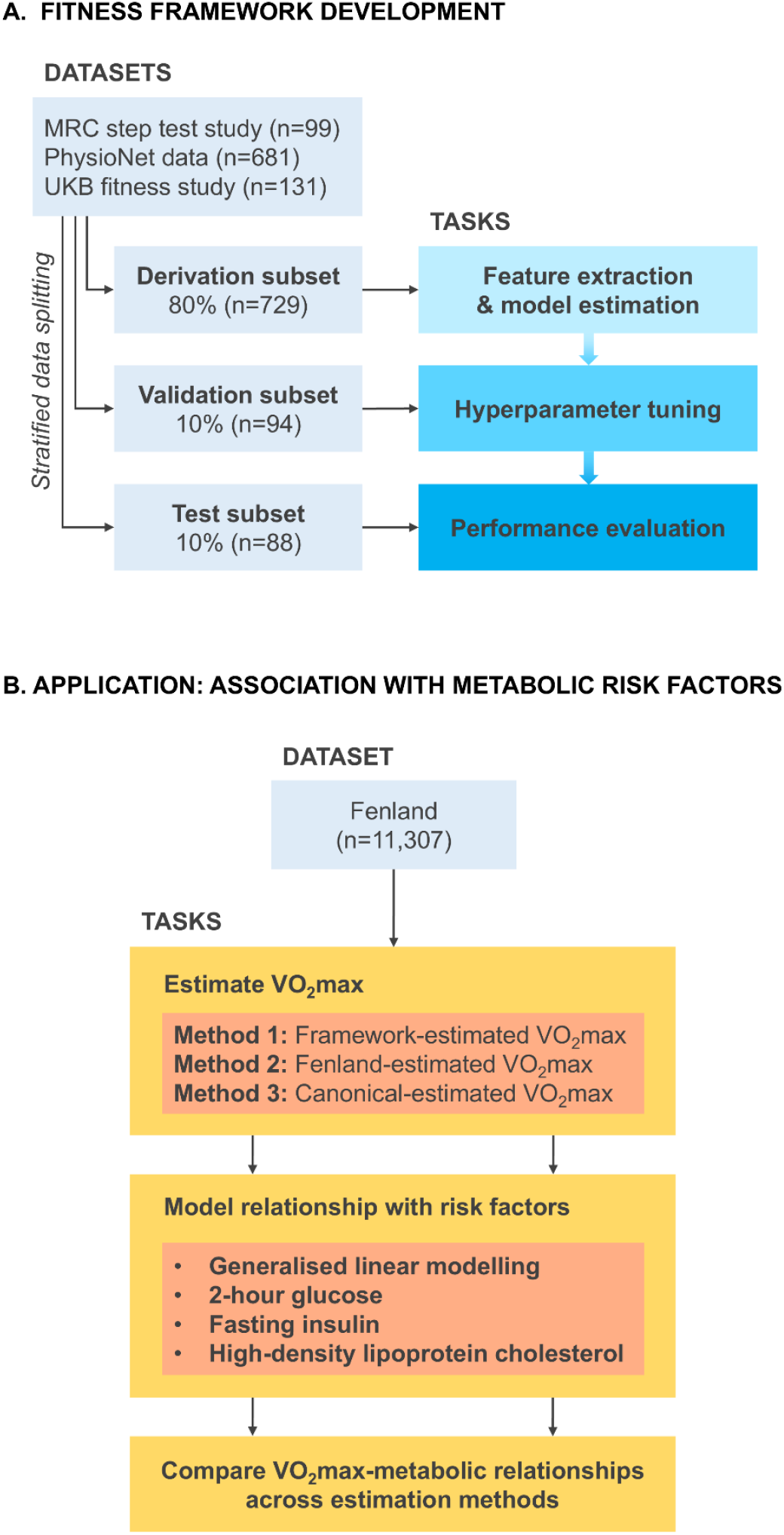
Flow diagram of the study design, comprising two phases: **A**. Development and evaluation of a novel framework to estimate 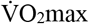 using exercise test data from several sources; and **B**. Application of the framework to examine associations between estimated 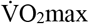 and metabolic risk in the Fenland Study. 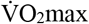 was estimated using the developed framework and two other approaches based on exercise heart rate response and predicted energy expenditure.

**Table 1.** Participant characteristics for datasets included in the framework development phase.

| Data subset | Derivation (n=729) |  |  |  |  | Validation (n=94) |  |  |  |  | Test (n=88) |  |  |  |  |
| --- | --- | --- | --- | --- | --- | --- | --- | --- | --- | --- | --- | --- | --- | --- | --- |
| Percentile | 5<sup>th</sup> | 25<sup>th</sup> | 50<sup>th</sup> | 75<sup>th</sup> | 95<sup>th</sup> | 5<sup>th</sup> | 25<sup>th</sup> | 50<sup>th</sup> | 75<sup>th</sup> | 95<sup>th</sup> | 5<sup>th</sup> | 25<sup>th</sup> | 50<sup>th</sup> | 75<sup>th</sup> | 95<sup>th</sup> |
| Age (y) | 19 | 25 | 33 | 44 | 60 | 18 | 26 | 32 | 46 | 59 | 20 | 26 | 32 | 47 | 61 |
| BMI (kg·m<sup>-2</sup>) | 20.4 | 22.6 | 24.2 | 25.9 | 30.0 | 19.9 | 22.8 | 24.1 | 25.5 | 29.2 | 20.5 | 22.5 | 24.1 | 26.5 | 29.8 |
| Weight (kg) | 56 | 67 | 74 | 82 | 95 | 57 | 64 | 73 | 79 | 96 | 55 | 66 | 71 | 84 | 97 |
| Height (cm) | 160 | 169 | 175 | 180 | 187 | 158 | 167 | 172 | 178 | 186 | 159 | 168 | 172 | 178 | 186 |
| Resting HR (bpm) | 54 | 60 | 61 | 63 | 69 | 50 | 60 | 61 | 63 | 68 | 56 | 61 | 62 | 63 | 72 |
| $\dot{V}O_2$max (ml O<sub>2</sub>·min<sup>-1</sup>·kg<sup>-1</sup>) | 27.4 | 37.7 | 44.2 | 50.8 | 58.5 | 25.9 | 38.1 | 44.7 | 51.7 | 58.3 | 26.2 | 37.2 | 44.1 | 51.7 | 58.9 |
| HRmax (bpm) | 161 | 176 | 183 | 190 | 200 | 157 | 174 | 183 | 189 | 201 | 157 | 172 | 181 | 191 | 200 |
| Sex (count) |  |  |  |  |  |  |  |  |  |  |  |  |  |  |  |
| Women | 23.2% (169) |  |  |  |  | 23.4% (22) |  |  |  |  | 22.7% (20) |  |  |  |  |
| Men | 76.8% (560) |  |  |  |  | 76.6% (72) |  |  |  |  | 77.3% (68) |  |  |  |  |

$\dot{V}O_2$ max: Maximal oxygen consumption, BMI: Body mass index, HR: Heart rate. HRmax: Maximal heart rate. Values are the 5<sup>th</sup>, 25<sup>th</sup>, 50<sup>th</sup>, 75<sup>th</sup>, and 95<sup>th</sup> percentiles, unless noted otherwise.

### Framework derivation, validation, and testing

The framework uses three interconnected models to generalise the relationship between the energetic demand of exercise and heart rate dynamics across a wide range of exercise modalities (see Methods for a detailed description). Briefly, the framework first standardises exercise intensity from different exercise modalities to oxygen consumption above resting levels 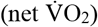. It then uses a linear model to relate net 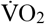 to corresponding target heart rate values, representing the heart rate expected for that level of exertion. Finally, in response to instantaneous changes in net 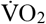, the framework simulates heart rate dynamics toward these target heart rate values. The framework’s simulation of heart rate dynamics is governed by individual-specific physiological parameters, including 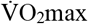, HRmax, and metabolic cost coefficients for different exercise modalities. We estimate these parameters for each participant by fitting their measured heart rate data to the framework’s simulated heart rate dynamics using Particle Swarm Optimisation (PSO). The PSO algorithm is guided by an objective function that balances two goals: 1) maximising agreement between simulated and measured heart rate, and 2) ensuring the physiological plausibility of parameter estimates. Physiological plausibility is ensured by minimising the Mahalanobis distance of the estimated parameters from a multivariate normal distribution (MVN) that captures expected physiological relationships based on individual characteristics like age, sex, height, and weight.

In the derivation dataset, we first fixed 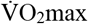, HRmax, and HRmin to directly-measured values and then used PSO to estimate the remaining framework parameters. These parameter values, along with individual-level characteristics, were then combined to estimate the sample MVN for computing Mahalanobis distances. **Figure 2A** displays the estimated mean vector and standard deviations (SD) of the MVN for men and women. Vector values were estimated in the log space and exponentiated to provide real-world values. Bounds for one SD above and below the mean were also calculated in log space and exponentiated. **Figure 2B** presents the unstandardised correlation matrix from the MVN, with cells displaying Pearson’s correlation coefficients. **Figure 2C** presents the standardised partial correlation matrix from the MVN, conditional on age, sex, weight, height, and minimum heart rate (HRmin), with cells displaying the partial correlation coefficients. The matrices reveal interrelationships among the framework parameters, and differences in the strengths of these relationships upon conditioning.

**Figure 2.**
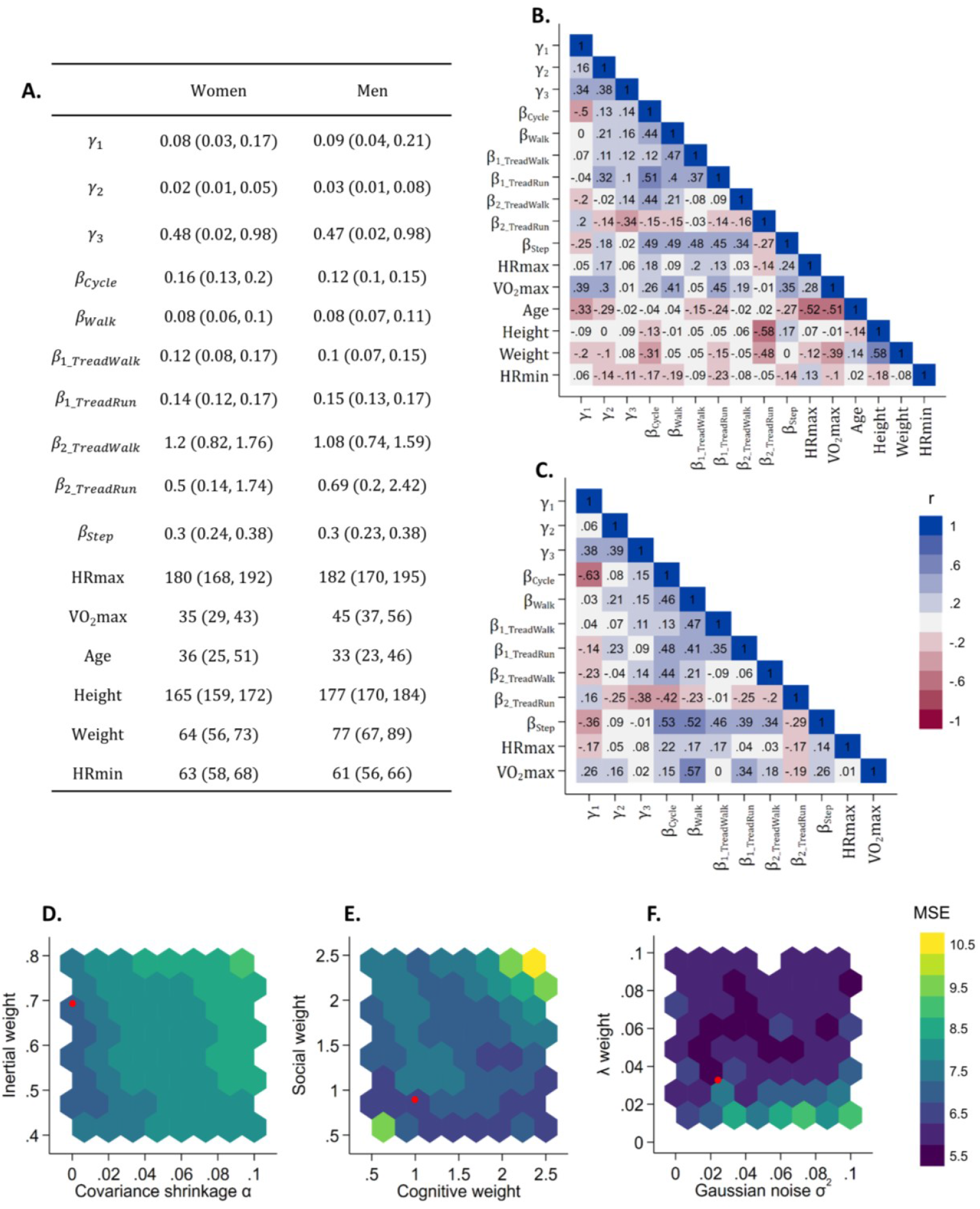
Characterisation and optimisation of the fitness framework. **A**. Mean vector and standard deviations (SD) of the multivariate normal distribution (MVN) for men and women. Vector values were estimated in the log space and exponentiated to provide real-world values. Bounds for one SD above and below the mean were also calculated in log space and exponentiated. **B**. Unstandardised correlation matrix from the MVN, with cells displaying Pearson’s correlation coefficients. **C**. Standardised partial correlation matrix from the MVN, conditional on age, height, weight, and minimum heart rate (HRmin), with cells displaying the partial correlation coefficients. **D-F**. Heat maps illustrating the relationship between hyperparameter values (inertial weight, social weight, cognitive weight, Mahalanobis distance penalisation weight *λ*, covariance shrinkage α, Gaussian noise σ) and 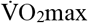 estimation mean squared error (MSE), derived from a heteroskedastic regression meta-model of framework performance. MSE values were predicted for each combination of hyperparameter values across Latin hypercube grid. Red dots represent hyperparameter values that minimise the predicted MSE from the meta-model. Optimised hyperparameter values were then used in the framework application phase of the study. Inertial weight: 0.693, Social weight: 0.895, Cognitive weight: 0.995, *λ*: 0.328, α: 0.000255, σ: 0.242.

With the sample MVN estimated, we next used the validation set to optimise parameter estimation for 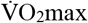. This involved a three-stage process to tune hyperparameters for PSO (inertial, social, and cognitive weights) and MVN regularisation (mean vector Gaussian noise, covariance shrinkage weight). First, we used PSO with Latin hypercube sampling to estimate 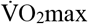 and other framework parameters in the validation set, exploring a wide range of hyperparameter combinations. Mahalanobis distances from the sample MVN guided the search process, ensuring physiological plausibility of parameter estimates. We then constructed a heteroskedastic regression meta-model to predict both 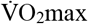 estimation bias and variance as a function of the hyperparameters and participant characteristics. Finally, we used gradient-based optimisation to minimise the average predicted mean squared error (MSE; squared estimation bias plus variance) from the meta-model, resulting in an optimal hyperparameter configuration. **Figure 2D-F** presents heatmaps illustrating the relationship between these hyperparameter values and the predicted MSE of 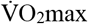 estimation. Red dots indicate the hyperparameter values that minimise the meta-model’s predicted MSE, which were subsequently used in the framework application phase of the study.

Having derived the sample MVN and optimised the hyperparameters, we then evaluated the framework’s performance in the independent test set, held out during derivation and parameter tuning. **Figure 3A** presents scatter plots and Bland-Altman plots illustrating agreement between framework-estimated and directly measured 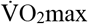 values. Overall mean estimation bias was low and correlation coefficients were strong (mean bias: 1.5 ml O_2_·min^-1^·kg^-1^; Pearson’s r: 0.80). Further agreement statistics are reported in **Supplementary Table 1**, as well as sensitivity analyses to evaluate the framework’s reliance on anthropometric information (height and weight) and close-to-max heart rate data (**Supplementary Figure 1**). This involved re-estimating 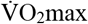 without conditioning the MVN on these variables and excluding some heart rate data during the PSO optimisation process. Restricting heart rate data to values below 85% of age-predicted maximum heart rate ^14^ slightly reduced agreement (mean bias: 2.0 ml O_2_·min^-1^·kg^-1^; Pearson’s r: 0.76). Further reductions in agreement were observed when also height and weight were removed (mean bias: 2.3 ml O_2_·min^-1^·kg^-1^; Pearson’s r: 0.69).

**Figure 3.**
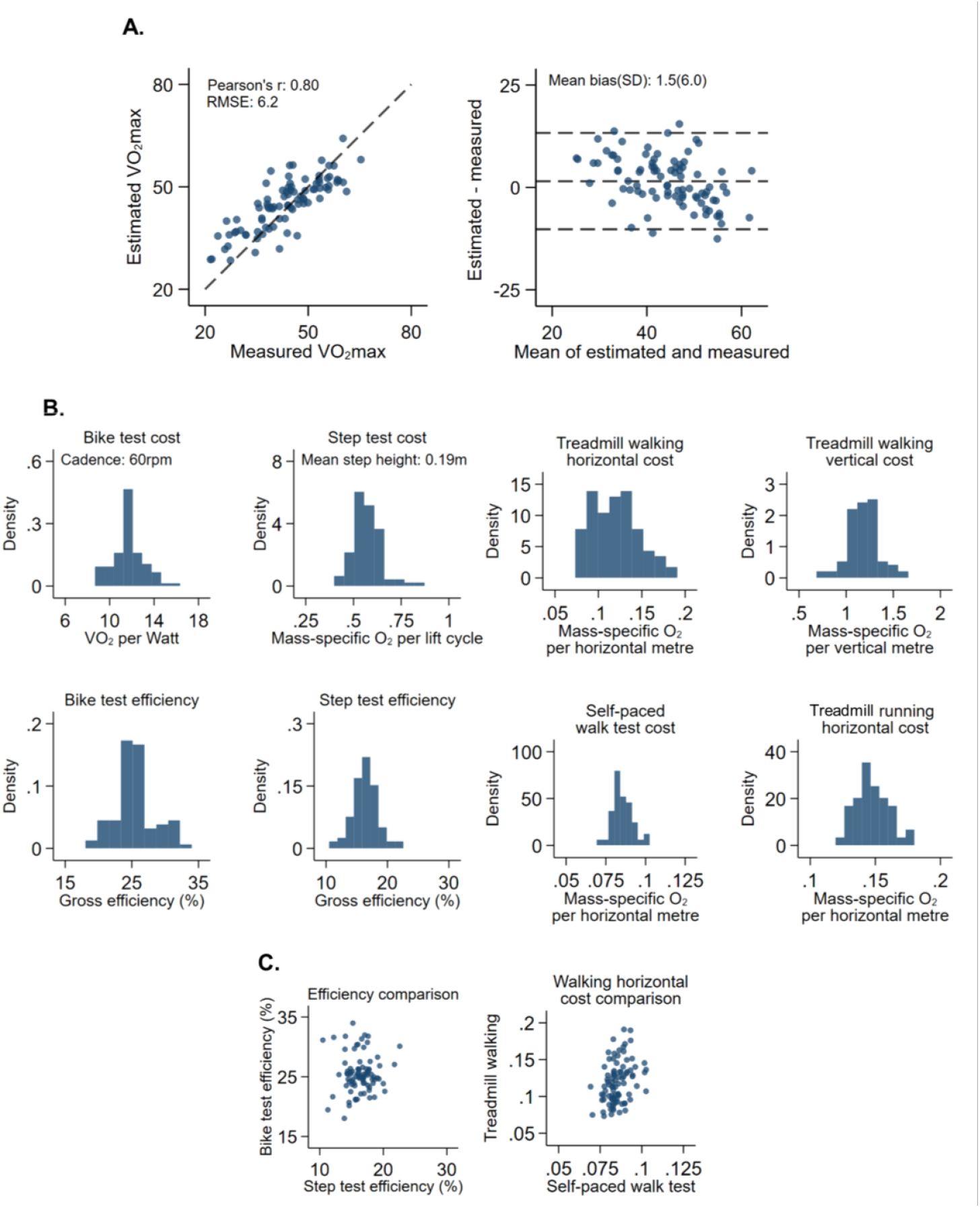
Performance evaluation of the fitness estimation. **A**. Scatter plots and Bland-Altman plots demonstrating agreement between framework-estimated and directly measured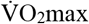. RMSE: Root mean squared error. **B**. Histograms of metabolic calibration coefficients estimated by the framework for different exercise modalities. Bike test and step test coefficients have been transformed to facilitate comparison with metabolic coefficients published by the American College of Sports Medicine (ACSM). **C**. Scatter plots comparing gross efficiency estimates for bike and step tests, as well as horizontal cost and treadmill and self-paced walking.

**Figure 3B** presents metabolic calibration coefficients estimated by the framework for different exercise modalities. Calibration coefficients were approximately normally distributed and generally similar to values published by ACSM (**Table 2**) ^15^. Estimated gross efficiency for cycle ergometry was 26 ± 3% and for step testing was 15 ± 2%, ranges that are within previously published gross efficiency values (**Figure 3C)**. The metabolic cost of treadmill walking was 25 ± 8% higher than self-paced walking when performed at similar speeds.

**Table 2.** Comparison of framework-estimated metabolic coefficients with American College of Sports Medicine (ACSM) published values.

| Metabolic coefficient | Equivalent ACSM published value | Marginal mean (95%CI) |
| --- | --- | --- |
| $\beta_{Walk}$ | n/a | 0.086 (0.084,0.087) |
| $\beta_{1TreadWalk}$ | 0.10 | 0.12 (0.096,0.14) |
| $\beta_{2TreadWalk}$ | 1.80 | 1.18 (1.09,1.28)** |
| $\beta_{1TreadRun}$ | 0.20 | 0.15 (0.14, 0.15)** |
| $\beta_{2TreadRun}$ | 0.90 | 0.75 (0.68,0.82)* |
| $\beta_{Cycle}$ | 11.1 | 11.7 (10.5,13.0) |
| $\beta_{Step}$ | 0.65 | 0.57 (0.47,0.67) |

$\beta$ : Exercise modality-specific metabolic coefficients estimated using the framework. Marginal mean values were computed from linear regression models adjusted for sex, age, and body mass index. ACSM: American College of Sports Medicine. \* ( $p < 0.05$ ) and \*\* ( $p < 0.01$ ) represent statistically significant difference from the corresponding ACSM published value.

### Examination of associations with cardiometabolic risk

To demonstrate its construct validity, we applied the framework to the Fenland Study cohort (**Table 3**) and found that framework-estimated 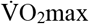 had stronger associations with key cardiometabolic risk factors compared to two conventional methods (Fenland-estimated and Canonical-estimated; **Table 4**). In models adjusted for age, sex, and resting heart rate, one-standard deviation (5.43 O_2_·min^-1^·kg^-1^) higher framework-estimated 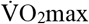 was associated with 10% lower 2-hour glucose, 37% lower fasting insulin, and 9.4% higher high-density lipoprotein (HDL) cholesterol. In contrast, one-standard deviation (11.2 ml O_2_·min^-1^·kg^-1^) higher canonical-estimated 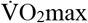 was associated with 4.4% lower 2-hour glucose, 12% lower fasting insulin, and 2.6% higher HDL. Fenland-estimated 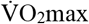 associations were between these two sets of estimates. A combined model of all three risk factors showed a substantially better statistical fit for framework-estimated fitness (AIC = 125,743) compared to Fenland-estimated (AIC = 128,849) and Canonical-estimated (AIC = 129,227) fitness. While further adjustment for height and weight attenuated all associations, the framework’s estimates retained the strongest associations with all outcomes (**Figure 4**). Median 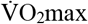 estimated by the framework was lower for both women (33.1 ml·min^-1^·kg^-1^) and men (39.4 ml·min^-1^·kg^-1^) compared to values from the Fenland-estimated (35.0 and 42.0 ml·min^-1^·kg^-1^) and Canonical-estimated (43.4 and 51.7 ml·min^-1^·kg^-1^) approaches.

**Figure 4.**
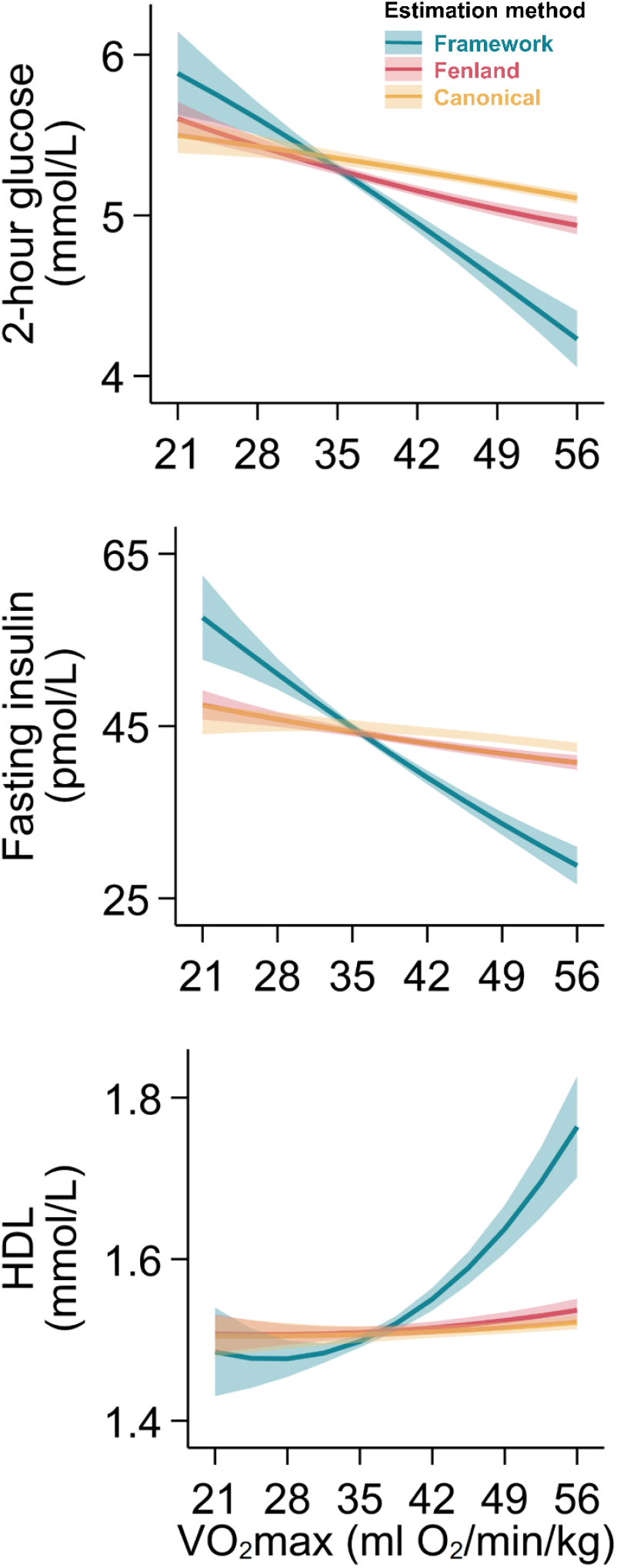
Associations of estimated 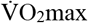 (ml O_2_·min^-1^·kg^-1^) with 2-hour glucose, fasting insulin, and high-density lipoprotein (HDL) cholesterol. Lines represent estimated marginal means and 95% confidence intervals for associations after adjustment for age, sex, resting heart rate, height, and weight.

**Table 3.** Fenland Study participant characteristics.

|  | Women (n=5,958) | Men (n=5,349) |
| --- | --- | --- |
| Sex |  |  |
| Age (y) | 48.2 (42.4-54.0) | 48.3 (42.2-54.3) |
| BMI (kg·m<sup>-2</sup>) | 25.2 (22.7-28.7) | 26.7 (24.4-29.3) |
| Weight (kg) | 68 (61-78) | 84.4 (76.4-93.4) |
| Height (cm) | 164 (160-168) | 178 (173-182) |
| Resting heart rate (bpm) | 64 (59-70) | 61 (55-67) |
| 2-hour glucose (mmol·L<sup>-1</sup>) | 5.0 (4.2-5.9) | 5.1 (4.2-6.1) |
| Fasting insulin (pmol·L<sup>-1</sup>) | 34.2 (24.2-49.9) | 42.8 (29.1-64.1) |
| HDL cholesterol (mmol·L<sup>-1</sup>) | 1.66 (1.41-1.94) | 1.30 (1.11-1.53) |
| Framework-estimated $\dot{V}O_2\text{max}$ (ml O<sub>2</sub>·min<sup>-1</sup>·kg<sup>-1</sup>) | 33.1 (30.6-35.9) | 39.4 (36.8-42.7) |
| Fenland-estimated $\dot{V}O_2\text{max}$ (ml O<sub>2</sub>·min<sup>-1</sup>·kg<sup>-1</sup>) | 35.0 (30.5-40.3) | 42.0 (37.7-49.0) |
| Canonical-estimated $\dot{V}O_2\text{max}$ (ml O<sub>2</sub>·min<sup>-1</sup>·kg<sup>-1</sup>) | 43.4 (37.4-49.2) | 51.7 (46.2-57.2) |

BMI: Body mass index, HDL: High-density lipoprotein cholesterol. Values are median (interquartile range), unless noted otherwise.

**Table 4.** Comparison of 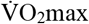 estimation methods and their association with cardiometabolic risk in the Fenland Study cohort. Coefficients represent the percent difference in each risk factor per 1-standard deviation (SD) difference in 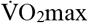 from generalised linear models (gamma distribution, log link). AIC values are presented as a measure of overall statistical fit across the three risk factors, allowing for a direct comparison between the framework, Fenland-estimated, and Canonical-estimated approaches within each adjustment model.

|  | Model 1 |  |  | Model 2 |  |  |
| --- | --- | --- | --- | --- | --- | --- |
| | Framework-estimated<br>$\dot{V}O_{2\max}$ | Fenland-estimated<br>$\dot{V}O_{2\max}$ | Canonical-estimate<br>$\dot{V}O_{2\max}$ | Framework-estimated<br>$\dot{V}O_{2\max}$ | Fenland-estimated<br>$\dot{V}O_{2\max}$ | Canonical-estimated<br>$\dot{V}O_{2\max}$ |
| 2-hour glucose | -0.10 (-0.11, -0.095) | -0.061 (-0.067, -0.054) | -0.044 (-0.050, -0.038) | -0.050 (-0.059, -0.040) | -0.034 (-0.041, -0.027) | -0.026 (-0.031, -0.020) |
| Fasting insulin | -0.37 (-0.38, -0.36) | -0.17 (-0.18, -0.15) | -0.12 (-0.13, -0.11) | -0.11 (-0.12, -0.087) | -0.041 (-0.053, -0.029) | -0.032 (-0.043, -0.022) |
| HDL cholesterol | 0.094 (0.088, 0.10) | 0.036 (0.031, 0.042) | 0.026 (0.021, 0.031) | 0.022 (0.014, 0.030) | 0.0057 (0.0031, 0.011) | 0.0058 (0.0010, 0.010) |
| AIC | 125,743 | 128,849 | 129,226 | 123,095 | 123,307 | 123,351 |

Model 1: Adjusted for age, sex, resting heart rate
Model 2: Adjusted for age, sex, resting heart rate, height, weight
AIC: Akaike's Information Criterion, HDL: High-density lipoprotein cholesterol. Reported values are semi-elastic beta coefficients and 95% confidence intervals for percent difference in each metabolic risk factor per 1-standard deviation (SD) difference in $\dot{V}O_{2\max}$ . Framework-estimated $\dot{V}O_{2\max}$ SD: 5.43 ml O<sub>2</sub>·min<sup>-1</sup>·kg<sup>-1</sup>. Fenland-estimated $\dot{V}O_{2\max}$ SD: 9.60 ml O<sub>2</sub>·min<sup>-1</sup>·kg<sup>-1</sup>. Canonical-estimated $\dot{V}O_{2\max}$ SD: 11.2 ml O<sub>2</sub>·min<sup>-1</sup>·kg<sup>-1</sup>.

## DISCUSSION

Here we present the development, evaluation, and application of a generalised framework for estimating cardiorespiratory fitness across a wide range of exercise testing modalities and protocols. To our knowledge, this approach is among the first that can be applied to the diversity of exercise tests commonly used in population health research, addressing the persistent challenge of method variability in this field.

The framework allows for individual variation of the metabolic cost of exercise, leading to more precise estimates of 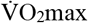 and other physiological parameters. For example, median metabolic cost coefficients estimated by the framework were closely aligned with group-level values published by the ACSM for treadmill walking, cycle ergometry, and step testing ^15^. This demonstrates that established principles of exercise physiology are an emergent property of the framework. The notable deviations observed for the ACSM’s coefficients were for the interaction of walking speed and treadmill gradient, as well as treadmill running. These deviations align with previous reports that have also questioned these specific ACSM coefficients ^16–18^. This suggests that our framework is not only valid but may even be more robust, as it independently identifies known weaknesses in current field-use equations. Taken together, these findings suggest the framework captures a more fundamental aspect of an individual’s exercise physiology from which established group-level metabolic cost coefficients can be derived.

This ability to capture a more fundamental layer of physiology translates directly into clinical utility, as demonstrated by the framework’s associations with cardiometabolic risk factors in the Fenland study. Statistically, framework-estimated 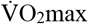 yielded markedly lower AIC values, indicating that it may explain substantially more variance in metabolic health than conventional methods. Physiologically, these statistics translate into steeper and more pronounced dose-response relationships for 2-hour glucose, fasting insulin, HDL cholesterol. This was observed even when associations were modelled as difference in risk factor per standard deviation differences in estimated 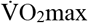 and adjusting for all covariates used to condition the framework. This performance likely stems from the framework’s ability to capture individual metabolic variation discussed previously, rather than relying on group-level assumptions inherent in conventional methods. These stronger associations have direct clinical implications, making applications such as personalised exercise prescriptions and more accurate tracking of intervention responses increasingly possible. This foundational work provides a validated and more sensitive tool for future etiological investigations to comprehensively elucidate complex fitness-health relationships, particularly those that would integrate multi-omics data ^19,20^.

When applied in the Fenland Study, 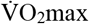 estimates from the framework were lower than those from methods that rely on linear extrapolation from steady-state assumptions. This is not unexpected and may, in fact, represent a more realistic estimation of the cohort’s fitness. Traditional methods often overestimate 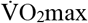 from ramped submaximal tests because participants may not achieve a true physiological steady state at different test stages. This leads to a heart rate-workload relationship which, when extrapolated to a presumed maximum heart rate, yields an inflated fitness estimate ^2^. This bias was less pronounced for the conventional model that used calibrated metabolic cost of the ramp protocol, thus lending support to this notion ^13^. By dynamically modelling the heart rate response without assuming steady state and with a less stringent assumption on maximum heart rate, the framework is designed to mitigate this source of overestimation bias.

Other possible applications for the framework include heart rate response to exercise protocols using both progressively increasing and decreasing workloads, as well as daily activities in free-living environments. The framework should be able to cope with mixed workload protocols, but no such data was available in our dataset. The intermittent and irregular nature of daily physical activity leads to complex heart rate and metabolic cost kinetics that may not be fully captured by our current model. The accuracy and precision of heart rate data from consumer wearables used in free-living settings may also be lower than research-grade monitors used in our study ^21^. Future research should evaluate and adapt the framework to these settings. This could include incorporating heuristics to better delineate true activity periods from sporadic movements and to identify and manage unreliable data segments from wearables. Comparisons with methods designed for passive fitness estimation in free-living environments will be crucial to determine the framework’s applicability in these contexts ^22^.

This study has limitations. Exercise test data used to derive and validate our framework predominantly originate from adults and excludes both children and adolescents. Although our framework is designed to be universally applicable, its conceptualisation as such would benefit by including data from a wide array of population and disease groups.

Future work should focus on validating the framework’s performance across diverse global populations. Additionally, exercise test data used to construct the framework were predominantly from those with higher fitness levels and contained a higher proportion of men compared to women. It is recognised that this analytic sample is not fully representative of the general population, which could potentially impact generalisability when the framework is applied to populations with lower fitness levels or higher proportions of women. The data underpinning the framework also does not have cycling at cadences other than 60 rpm and step test data from different step heights from the same individuals. In terms of framework parameter estimation, we used PSO; alternative optimisation methods, such as simulated annealing ^23^, genetic algorithms ^24^, ant colony optimisation ^25^, or a hybrid of these methods, may provide improvements to computational efficiency or scalability. Finally, while the parameters of the dynamical heart rate model have clear mathematical utility, elucidating their precise physiological basis remains an important avenue for future research.

We have presented and validated a generalised framework for estimating fitness from exercise heart rate response. The framework unifies current fitness estimation approaches, offering a versatile and adaptable way to estimate fitness from a wide range of exercise testing modalities and protocols. We demonstrate its application in estimating fitness in a large-scale population study to examine associations with cardiometabolic risk factors, yielding strong relationships compared to other approaches. By providing a test-agnostic method for estimating fitness, the framework is positioned to resolve the long-standing challenge of method variability, ultimately contributing to more inclusive and robust approaches for assessing fitness in both clinical and population health research.

## METHODS

### Exercise test data sources

This study used data from four sources, comprising three for framework development (Biobank validation study, Step test validation study, and a PhysioNet repository) and one for framework application (the Fenland Study). The development datasets included a variety of submaximal exercise tests, such as treadmill, cycle ergometer, self-paced walking, and step tests, and maximal exercise tests with directly measured 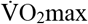. The application datasets (Fenland) used a submaximal treadmill protocol. Each study had distinct participant recruitment profiles and fitness assessment procedures, which are described in detail in their respective publications and protocol documents. Here we provide a summary of participant inclusion criteria, testing methods, and ethics approval for each study.

#### Biobank validation study

Data from the Biobank validation study were originally used to validate fitness and physical activity assessments in UK Biobank ^2,26^. Participants completed six submaximal exercise tests and a maximal test over two clinical visits and a free-living period. All assessments were completed within a two-week testing period. The clinical tests consisted of a 200-meter self-paced walk test, a ramped treadmill test, two ramped cycle ergometer tests, a steady-state cycle ergometer test, and maximal cycle ergometer test. The maximal test immediately followed the steady-state test, was the last test performed on a given visit, and was performed to directly measure 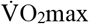 and maximal heart rate. During the free-living period, participants completed a ramped step test in their home environment. Exclusion criteria were: 1) heart pacemaker; unable to walk without aid; 2) history of angina pectoris; 3) blood pressure greater than 180/110 mm Hg; 4) musculoskeletal injury that would impair cycling on the ergometer; pregnancy; and 5) currently taking cardioactive drugs (e.g. beta-blockers, aspirin). Ethical approval was obtained by the University of Cambridge Human Biology Research Ethics Committee (Ref: HBREC/2015.16).

#### Step test validation study

Data from the step test validation study ^27^ were previously used to validate the Cambridge Ramped Treadmill Test from the Fenland study ^13,28^. The study recruited 51 young adults and 48 older adults from an existing database for research volunteers. Inclusion criteria were healthy women and men able to walk independently. Participants completed the step test and a maximal treadmill test during a clinical visit. The step test protocol consisted of body lifts (one body lift represents a full step-up and step-down cycle) performed on a 15 or 20 cm step over the following stages: 1) 1 minute of body lifts at a rate of 15 lifts per minute; 2) 7 minutes of body lifts with the rate increasing linearly from 15 to 32.5 lifts per minute; and 3) 2 minutes seated to measure recovery heart rate. Exclusion criteria were: 1) contraindications to exercise as measured by the Physical Activity Readiness Questionnaire; 2) any undiagnosed heart condition as determined by resting electrocardiogram (ECG); 3) self-reported breathlessness during limited stair climbing or walking; 4) pregnancy; 5) current reported use of cardioactive medications such as beta blockers; and 6) systolic and diastolic blood pressure greater than 180/110 mmHg. Ethical approval was obtained by the Huntingdon & Peterborough/Fenland Research Ethics Committee (Reg: 07/Q0106/21).

#### Exercise tests from the University of Malaga Exercise Physiology Lab

We used publicly-available maximal treadmill test data provided by the PhysioNet repository, which is managed by the Massachusetts Institute of Technology Laboratory for Computational Physiology ^29^. The database contained 992 maximal treadmill tests conducted from 2005 to 2018 in the Exercise Physiology and Human performance Lab of the University of Malaga ^30,31^. Data were used in the present analysis in accordance with the access policy detailed in the PhysioNet Contributor Review Health Data License 1.5.0 ^32^. Treadmill test data consisted of a mixture of steady-state and ramped submaximal and maximal protocols, including level walking, graded walking, and running. Heart rate, pulmonary gas exchange, and exercise intensity were measured during each treadmill test. Tests were completed by amateur and professional athletes aged 10 to 63 years old. Only valid exercise test data performed by adults were used in the present study (n=685).

#### Fenland Study

The Fenland Study is a prospective cohort study that recruited 12,435 participants born between 1950 and 1975. Participants were recruited from general practice lists around Cambridgeshire, UK, between January 2005 and April 2015 ^12^. Individuals with prevalent diabetes, pregnant or breastfeeding women, those unable to walk unaided for at least 10 minutes, individuals with psychosis, or those with a terminal illness were excluded. 5,958 women and 5,349 men were included in this analysis after exclusions (**Table 3**). Participants performed a submaximal treadmill test consisting of four stages: 1) 3 minutes of walking at 3.2 km·h^-1^ and 0% incline; 2) 6 minutes of walking with speed increasing from 3.2 to 5.2 km·h^-1^ at 0% incline; 3) 3 minutes of walking at 5.2 km·h^-1^ while incline increased from 0 to 6%, followed by 3 minutes of walking with speed increasing from 5.2 to 5. km·h^-1^ and incline increasing from 6 to 10.2%; and 4) 1 minute of running with speed increasing from 5.8 to 9.0 km·h^-1^ while incline decreased to 0%, followed by 4.5 minutes of running with speed increasing from 9.0 to 12.6 km·h^-1^ at 0% incline ^33^. HR data were recorded continuously using a chest-worn HR monitor ^34^. Cardiometabolic health was assessed via blood samples drawn following an overnight fast and 2 hours post glycaemic load ^35^. Here we report associations with 2-hour glucose, fasting insulin, and HDL cholesterol. Ethical approval was obtained from the Health Research Authority NRES Committee East of England-Cambridge Central. All participants provided written informed consent. A more detailed description of fitness in the Fenland Study can be found elsewhere ^13^.

### Framework architecture

A flow diagram describing the main components of the framework is shown in **Supplementary Figure 2**. The framework takes as input exercise heart rate data and a corresponding, time-varying measure of exercise intensity (e.g., walking speed, treadmill speed and grade, cycling power, stepping frequency and step height). In the present analysis, heart rate and exercise intensity data were interpolated to 1-second resolution, unless they were already recorded at that resolution. No further processing steps were applied to heart rate data, which were already pre-processed using methodologies specific to their original studies.

#### Calibration of exercise intensity to net energy expenditure

Group-level metabolic equations can yield imprecise energy expenditure estimates when applied to individual-level exercise test data ^33,36^. To address this issue, the first component of the framework uses a system of metabolic equations to individually calibrate exercise intensity from different exercise modalities (e.g. treadmill, cycle ergometer) to net energy expenditure (net VO_2_), defined as energy above resting energy expenditure (**Supplementary Figure 3A and 3B**). This approach is similar in principle to previous work that harmonised expressions of energy expenditure across different exercise modalities ^33^.

The system of equations used in this study was:

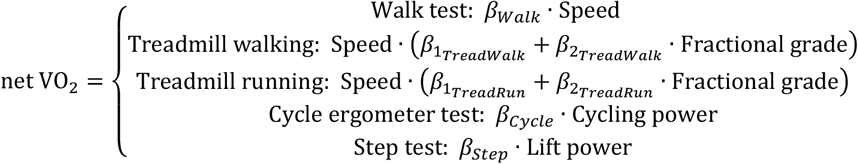

where each equation corresponds to the individualised energetic cost of the indicated exercise modality, and:

- ‘net VO_2_’ is oxygen consumption scaled by total-body mass (ml O_2_·min^-1^·kg^-1^)
- ‘Cycling power’ is in watts (W)
- ‘Speed’ is in metres·min^-1^
- ‘Fractional grade’ is treadmill incline percentage expressed in decimal form
- ‘Lift power’ is in J·min^-1^·kg^-1^, computed as the product of step frequency (lifts·min^-1^), step height (metres·lift^-1^), and gravitational acceleration (9.81 metres·sec^-2^) ^37^

The functional forms of the equations nested within net VO_2_ were derived from group-level metabolic equations published by the American College of Sports Medicine (ACSM) ^15^. Rather than apply the ACSM group-level calibration coefficients, the framework instead individualises this calibration, represented here as beta coefficients (*β*). This approach allows the framework to account for between- and within-individual differences in the metabolic cost and efficiency of different exercise modalities. We assumed that individual calibration would yield *β* coefficients that generally align with ACSM’s published group-level values. We examine this assumption later in the present analysis (see ‘Assessment of metabolic coefficients’). The derived units of measurement for each *β* coefficient were:

- *β*_*Walk*_: Mass-specific O_2_ cost per horizontal metre (ml O_2_·metre^-1^·kg^-1^)
- 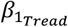 : Mass-specific O_2_ cost per horizontal metre (ml O_2_·metre^-1^·kg^-1^)
- 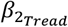: Mass-specific O_2_ cost per vertical metre (ml O_2_·metre^-1^·kg^-1^)
- *β*_*Cycle*_: 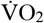per Watt (ml O_2_·min^-1^·kg^-1^·W ^-1^)
- *β*_*Step*_: O_2_ cost per unit of mechanical work (ml O_2_·J^-1^)

We specified separate metabolic equations for treadmill walking and treadmill running to account for within-individual differences in the energetic cost of these exercise modalities ^38^. The treadmill walking equation was applied when treadmill speeds were less than or equal to 90 metres·min^-1^. The treadmill running equation was applied when treadmill speeds were greater than or equal to 120 metres·min^-1^. These thresholds generally delineate the transition from walking to running in adult human locomotion ^39,40^.

The transition from walking to running (e.g., between 90 and 120 metres·min^-1^) was modelled using the following linear interpolation equation for treadmill speed:

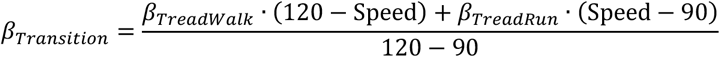

where *β*_*Transition*_ is either *β*_1_ or *β*_2_ in the treadmill test equation and ‘Speed’ is treadmill speed. This approach enables the energetic cost of treadmill exercise to be individually calibrated across many potential testing protocols.

#### Calibration of net oxygen consumption to target heart rate

Heart rate increases linearly with oxygen consumption and reaches a maximal value when 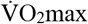 is achieved ^41^. Based on this principle, the framework uses the following linear equation to calibrate net VO_2_ values to ‘target’ heart rate values (HR_target_):

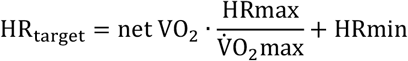

where:

- HRmin is minimal heart rate
- HRmax is maximal heart rate
- 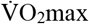 is maximal oxygen consumption scaled by total-body mass (ml O_2_·min^-1^·kg^-1^)
- HR_target_ are heart rate values that correspond to different energy expenditure levels

The ‘HRmax to 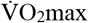 ratio’ can be conceptualised as the oxygen pulse at maximal exercise. The ratio’s value anchors the *β* coefficients for calibrating exercise intensity to net VO_2_ to values that would allow heart rate and oxygen uptake kinetics to be aligned.

#### Modelling of heart rate dynamics

Exercise heart rate response kinetics have historically been modelled using multiple mono-exponential functions for growth and decay ^42^. However, within-individual, the time constants for these functions can vary widely when applied to different exercise intensity profiles, making them difficult to interpret across exercise tests ^43^. We propose a model of heart rate dynamics where onset and offset heart rate kinetics emerge from the model’s behaviour, rather than functions with fixed time constants (**Supplementary Figure 3C**). After calibrating oxygen consumption to corresponding HR_target_ values, we use a series of coupled differential equations to model heart rate dynamics across the physiological intensity spectrum:

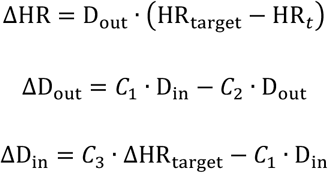

The first equation models change in heart rate (ΔHR) at time *t* (HR_*t*_) as the difference between HR_target_ for a given net VO_2_ value and the current heart rate value multiplied by an ‘output drive’ parameter (D_out_). The parameter D_out_ controls the rate at which instantaneous heart rate approaches a target heart rate value. The second equation uses two factors to model change in the ‘output drive’ parameter (ΔD_out_): a positive ‘input drive’ parameter (D_in_) scaled by *C*_1_ and a negative factor that decays the current value of D_out_ by *C*_2_. Change in the ‘input drive’ parameter (ΔD_in_) is also modelled using two factors: a positive factor proportional to ΔHR_target_ scaled by *C*_3_ and a negative factor that decays the current value of D_in_ by *C*_1_. The positive and negative factors in the second and third equations (*C*_1_ · D_in_) are equivalent in magnitude.

The values of the scaling parameters (*C*_1_, *C*_2_, *C*_3_) within the series of differential equations change dynamically with exercise intensity. These values are determined by linear functions that relate the current heart rate to heart rate reserve:

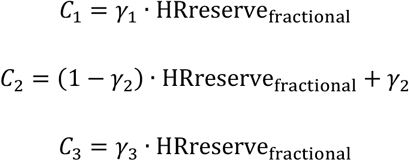

where *γ*_1_, *γ*_2_, and *γ*_3_ are estimated coefficients defining the slope of their respective linear functions and HRr_fractional_ is fractional heart rate reserve:

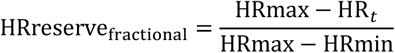

HRreserve_fractional_ is a normalised, inverse representation of exercise intensity that ranges from 1 (minimal intensity, HR=HRmin) to 0 (maximal intensity, HR=HRmax). When estimated by the framework, *γ*_1_ and *γ*_2_ are bound within the interval 0 to 1 and *γ*_3_ is strictly positive.

The proposed heart rate dynamics model controls input drive signals caused by changes in energy expenditure during exercise and operating within physiological constraints. This process dynamically regulates the heart’s kinetic profile when changing to a new target heart rate. For example, when approaching the heart’s maximal ventricular beating rate, the model prevents instantaneous heart rate from exceeding this value. Instantaneous heart rate heart is also prevented from falling below a minimal value.

### Estimation of model parameters

The framework consists of three interconnected models, each with its own set of parameters:

- Calibration of exercise intensity to net energy expenditure: *β*_*Walk*_, 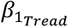, 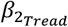, *β*_*Cycle*_, *β*_*Step*_
- Calibration of net energy expenditure to target heart rate: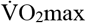, HRmax, HRmin
- Heart rate dynamics: *γ*_1_, *γ*_2_, *γ*_3_

The goal of parameter estimation is to identify the set of parameter values that best fits the framework’s simulation of heart rate dynamics to measured exercise heart rate data. This process involves these heart rate measurements, exercise intensity data, and individual-level characteristics to estimate the parameters listed above.

### Parameter estimation using particle swarm optimisation

Due to the novel architecture of the framework, we used Particle Swarm Optimisation (PSO), a metaheuristic algorithm for complex optimisation problems (**Supplementary Figure 4**) ^44^. PSO guides the behaviour of a “swarm” of particles, where each particle represents a vector of parameter values intended to optimise the model. Particles explore the parameter space and iteratively adjust their trajectories based on a specified objective function (described in detail below). By sharing information about their positions and performance with other particles, the swarm gradually converge towards an optimal set of parameters. The particle that achieves the best objective function value upon termination of the algorithm is selected as the final estimated parameter set.

In our implementation, each PSO instance used 64 particles. Initial parameter values for each particle were randomly sampled from uniform distributions within predefined ranges (**Supplementary Table 2**). The algorithm was terminated after 64 iterations, a value chosen to balance computational efficiency with sufficient exploration of the parameter space. Particle movement and search behaviour were controlled by three hyperparameters: inertial weight, social weight, and cognitive weight. These hyperparameters control the influence of a particle’s own best position (determined by its objective function value) and the swarm’s best position on the particle’s trajectory. The values for these hyperparameters were determined as described in section “Tuning of framework hyperparameters”. A ring topology was used for the communication architecture, with each particle sharing its position with its two nearest neighbours. This topology was chosen because it promotes exploration of the parameter space and helps prevent premature convergence to a suboptimal solution ^45^.

#### Objective function with Mahalanobis distance penalty for physiological plausibility

We specified the PSO objective function, denoted *f*(*x*), to find a particle parameter vector *x* that satisfies two criteria: 1) agreement between modelled and observed heart rate data; and 2) adherence to physiological constraints. This is achieved through a two-component objective function. The first component maximises a composite coefficient of determination:

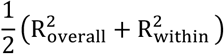

where:

- 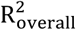 is the overall coefficient of determination, which quantifies the proportion of variance in the measured heart rate response explained by the modelled response
- 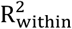 is the within-exercise-intensity coefficient of determination, which quantifies agreement between measured and modelled heart rate response at each distinct ‘target’ heart rate level

If only 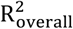 is used, the shapes of the measured and modelled heart rate response do not agree. Using only 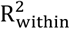 results in an offset between modelled and measured heart rate responses, despite their shapes agreeing. Defining the first component as the average of 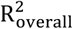 and 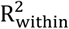 results in good overall agreement and shape correspondence between the measured and modelled heart rate responses.

While the first component of the objective function ensures a good fit to observed heart rate data, it does not guarantee that the resulting model parameters are always physiologically plausible. To address this, the objective function incorporates a penalty term that encourages solutions consistent with known physiological relationships between model parameters and individual characteristics. For example, younger, normal-weight adults tend to exhibit higher 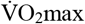 values and faster heart rate kinetics compared to older, obese adults. The penalty term is based on the Mahalanobis distance, denoted *d*_*M*_, which measures how far a given solution deviates from expected physiological norms (**Supplementary Figure 5**). Assuming these parameters and characteristics are jointly normally distributed in the population, *d*_*M*_ is computed using an underlying population multivariate normal (MVN) distribution, denoted 𝒩(*μ*, Σ):

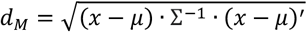

where:

- *d*_*M*_ is the Mahalanobis distance between a PSO particle parameter vector (*x*) from expected physiological norms, conditioned on individual-level characteristics
- *μ* is the population mean vector of the MVN, containing model parameters and individual-level characteristics
- Σ is the population covariance matrix of the MVN, describing interrelationships between framework parameters and individual-level characteristics.

The penalty term in the objective function is then defined as:

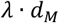

where *λ* is the physiological plausibility weight hyperparameter that controls the influence of *d*_*M*_ on the optimisation process. Combining the first component of the objective function and the penalty term yields the following overall objective function *f*(*x*):

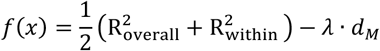

Defining the objective function as *f*(*x*) encourages the PSO to find solutions that maximise agreement between measured and modelled heart rate data while minimising the Mahalanobis distance from expected physiological norms. Appropriate tuning of *λ* is required to balance the trade-off between model fit and physiological plausibility.

Using the Mahalanobis distance, *d*_*M*_, as a penalty term allows for the estimation of individual-calibration parameters (*β*), even when data for a specific exercise modality are missing (i.e., the individual did not perform that exercise). While each PSO particle represents a complete set of parameters for all modalities, *d*_*M*_ is calculated using only the parameters corresponding to modalities performed by a given individual. However, because the MVN distribution encodes relationships between individual-level characteristics and all parameters, minimising *d*_*M*_ effectively constrains the values of unobserved parameters by conditioning them on observed parameters through the MVN’s covariance structure. In this way, prior information encoded in the MVN distribution is used to inform the estimation of parameters for which data are unavailable.

#### Regularisation of Mahalanobis distance penalty

In practice, the underlying population MVN 𝒩(*μ*, Σ) is unknown. Its parameters *μ* and Σ are instead estimated from a sample, yielding a sample-estimated MVN 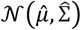 with sample mean vector 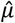 and sample covariance matrix 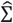. These are used to compute the Mahalanobis distance *d*_*M*_ in lieu of the population MVN:

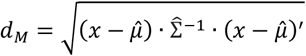

However, a sample-estimated MVN may be misspecified and overfit to the data it was derived from, leading to poor generalisation and overconfident estimates of *d*_*M*_. To mitigate the risk of the PSO overfitting to potentially spurious characteristics of a sample, we regularise the sample-estimated MVN by applying Gaussian noise to elements in 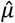 that correspond to individual-level characteristics:

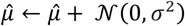

where:

- 𝒩(0, σ^2^) is zero-mean Gaussian noise with variance hyperparameter σ^2^

Furthermore, we also regularise the sample-estimated MVN by applying covariance shrinkage to 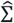:

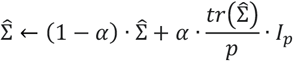

where:

- α is the shrinkage intensity hyperparameter
- *tr*(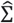) is the trace of 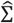
- *p* is the number dimensions in 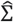
- *I*_*p*_ is the *p* x *p* identity matrix

Adding Gaussian noise to the individual-level characteristics in 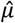 introduces uncertainty. This reduces the risk of overfitting to potentially sample-specific characteristics. Covariance shrinkage pulls 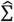 towards a scaled identity matrix, which has two primary benefits ^46^. First, it dampens the strength of estimated associations between parameters, reducing the influence of spurious correlations on *d*_*M*_ calculations. Second, it preserves the positive-definiteness of 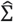, a necessary property for calculating *d*_*M*_. Appropriate tuning of σ^2^ and α are required to balance the benefits of regularisation while retaining sufficient information within the sample-estimated MVN.

### Model training

Each dataset (e.g. Biobank validation study, Step test validation study, PhysioNet treadmill exercise dataset) was split into derivation (80%), validation (10%), and test (10%) subsets using stratified sampling by age, weight, and sex. Derivation, validation, and test subsets were then pooled, respectively, for model training and evaluation.

#### Estimation of sample covariance matrix for Mahalanobis distance calculations

In the derivation set, we first fixed 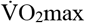, HRmax, and HRmin to their directly measured values. This ensured that relationships between these parameters, other estimated model parameters, and individual-level characteristics would be accurately reflected in the estimated sample MVN. We then used PSO to estimate values for all other framework parameters. For this process, PSO hyperparameters for inertial and social weight were each set to 1.3 and cognitive weight was set to 0.6. Only the first component of the objective function was applied, as the sample MVN had not been estimated thus far and could not be used to compute *d*_*M*_. Following this process, we defined a parameter vector for each participant that included the estimated parameters, directly measured values for 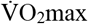, HRmax, and HRmin, and individual-level characteristics related to fitness (age, height, weight, sex). The parameter vectors were then combined into a single dataset.

To estimate the sample MVN, we applied the maximum-likelihood expectation-maximisation (EM) algorithm ^47^ to the combined dataset. We chose EM because it allowed us to utilise all participant data, including those missing parameters because of pooling datasets with different exercise testing protocols. We acknowledge that applying EM in this context violates the missing at random assumption, as the missingness is related to study origin. This could potentially introduce bias into the sample MVN. However, we opted for EM over a complete-case analysis to maximise the use of available data, especially given the regularization techniques described earlier that would be applied to mitigate potential bias in the sample MVN.

#### Tuning of framework hyperparameters

We used a three-stage hyperparameter optimization strategy to maximise agreement between estimated and directly-measured 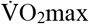. First, using the sample MVN from the derivation set, we applied PSO to estimate framework parameters in the validation set across a dense grid of hyperparameter value combinations. Grids were generated using Latin hypercube sampling, a space-filling design that efficiently explores the parameter space ^48^. For each participant, 512 unique hyperparameter combinations were generated, and three replicate estimations were performed for each combination to account for the stochastic nature of the PSO algorithm. We then evaluated framework performance for each combination as the difference between estimated and directly-measured 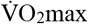 (e.g. estimation bias). This yielded a dataset of relationships between hyperparameters, participant characteristics, and framework performance.

Next, we constructed a heteroskedastic regression meta-model to predict both estimation bias and its variance as a function of the hyperparameters and participant characteristics. The meta-model served as a computationally efficient surrogate for the complex relationships between hyperparameters and model performance, allowing us to explore a wider range of hyperparameter values without repeatedly running the full PSO estimation. Importantly, heteroskedastic regression enables modelling of non-constant variance in estimation bias across different hyperparameter configurations, allowing the Mean Squared Error (MSE) of the framework to be computed directly. This allows for the identification of hyperparameters that optimally balance bias and variance, minimising the overall error. To capture potential non-linearities and heterogeneity in the bias-hyperparameter relationships, the meta-model incorporated main effects and all two-way interactions between hyperparameters as well as interactions with participant age and BMI.

Finally, to identify the optimal hyperparameter values, we defined a meta-objective function that calculated the average predicted MSE from the meta-model across a grid of age and BMI values. Newton-Raphson’s method was then used to minimise the meta-objective function, resulting in a hyperparameter configuration that minimised the average MSE of the framework across a range of individual characteristics.

### Model evaluation

#### Assessment of agreement

Using the sample MVN from the derivation set and optimal hyperparameter values identified in the validation set, we applied the framework to the test set to estimate both 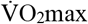 and metabolic cost coefficients. These values were estimated by fitting each individual’s measured heart rate to the framework’s simulated heart rate dynamics, as described previously. To evaluate agreement between framework-estimated and directly-measured 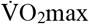, we used scatter plots and Bland-Altman plots and computed the following metrics:

- Mean bias error 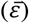, calculated as the average difference between measured and estimated values.
- Standard deviation of the mean error (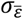).
- Root mean squared error (RMSE, calculated as 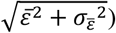
- Pearson’s correlation coefficient (*r*)
- Concordance correlation coefficient (*ρ*_*c*_), quantifies agreement by considering both correlation and bias. ^49^
- Bias correction factor (*C*_*b*_, calculated as *ρ*_*c*_⁄*r*), quantifies the deviation of the best-fit line from the line of perfect concordance (45° line). *C*_*b*_ values closer to 1 indicate minimal bias.

As a sensitivity analysis, we reapplied the framework to the test set under different data restriction scenarios: when not using weight and height to condition the MVN during parameter estimation, and when censoring data greater than 85% age-predicted maximal heart rate. These scenarios were explored to examine the framework’s reliance on both anthropometric data and heart rate data closer to maximal values.

#### Assessment of metabolic coefficients

We examined the assumption that the framework would yield individual-calibration parameters (*β* coefficients) that correspond to group-calibration coefficients published by ACSM. We used regression analysis to summarise *β* coefficient estimates for each exercise modality and compared their marginal mean values with ACSM’s group-level calibration coefficients ^15^. For reference, the ACSM coefficients are:

- Treadmill walking (horizontal component): 0.1 ml O_2_·metre^-1^·kg^-1^
- Treadmill walking (vertical component): 1.8 ml O_2_·metre^-1^·kg^-1^
- Treadmill running (horizontal component): 0.2 ml O_2_·metre^-1^·kg^-1^
- Treadmill running (vertical component): 0.9 ml O_2_·metre^-1^·kg^-1^
- Stepping (0.19 metre step height per lift cycle): 0.65 ml O_2_·lift^-1^·kg^-1^
- Cycle ergometry (60 rpm cadence): 11.1 ml O_2_·min^-1^·W^-1^

To facilitate comparison with the ACSM cycle ergometry coefficient, *β*_*Cycle*_ (ml O_2_·min^-1^·kg^-1^·W^-1^) was multiplied by total-body mass. Similarly, *β*_*Step*_ (ml O_2_·J^-1^) was multiplied by gravitational acceleration (9.81 metres·sec^-2^) and step height per lift cycle (metres·lift^-1^) to facilitate comparison with the ACSM stepping coefficient. The mean (range) step height used for step testing in the present analysis was 19 (15-20) cm. ACSM does not publish a metabolic equation for self-paced walking, but the metabolic cost of treadmill walking is reported to be greater than self-paced walking ^50^. We compared estimated *β*_*Walk*_ with *β*_*TreedWalk*_ to examine the percent difference in metabolic cost between modalities.

Since *β*_*Cycle*_ and *β*_*Step*_ each incorporate direct measurements of mechanical work, gross efficiency can be estimated for cycle ergometry and step testing. Gross efficiency is the ratio of mechanical work produced during exercise to the total metabolic energy expenditure required to produce that work. Assuming a caloric equivalent of oxygen of 20.35 J·ml O_2_^-1, 51^ gross efficiency for cycle ergometry (*E*_*Cycle*_) was computed as:

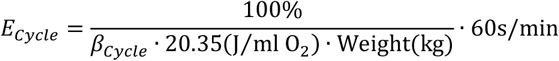

where ‘Weight’ is total-body mass. Gross efficiency for step testing (*E*_*Step*_) was computed as:

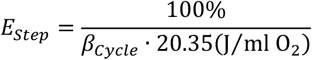

Gross efficiency values for cycle ergometry and step testing were compared within-individual and with previously reported ranges ^52,53^. When performing cycle ergometry at a fixed 60 rpm cadence, *E*_*Cycle*_ can range from 20% to 30%. *E*_*Step*_ can range from 10% to 20% at a step height of approximately 0.2 metres.

### Comparative analysis of associations with cardiometabolic risk

To assess the clinical utility of our framework and compare its performance against established methods, we estimated 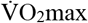 from exercise test data in the Fenland Study using three methods: 1) our novel framework; 2) a study-specific linear regression method previously developed for the Fenland study (Fenland-estimated) ^28^; and 3) a canonical linear regression method based on ACSM equations (Canonical-estimated) ^15^. Both the Fenland-estimated and Canonical-estimated approaches extrapolate the linear relationship between exercise heart rate response and exercise intensity to age-predicted maximal HR to estimate 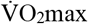 ^41^. We then examined associations between each 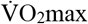 estimation method and three cardiometabolic risk factors: 2-hour glucose, fasting insulin, and high-density lipoprotein (HDL) cholesterol. These were chosen because they are indicators of distinct cardiometabolic health pathways (e.g., glycaemic control, insulin resistance, lipid metabolism) with known relationships with fitness ^54,55^.

We used a structural equation modeling (SEM) approach to simultaneously estimate and compare the association between each fitness estimate and the three risk factors. This approach allows for the evaluation of each fitness measure against the complete metabolic risk profile, providing a single measure of model fit (Akaike Information Criterion; AIC) for direct comparison. Within the SEM, the relationship for each risk factor was specified by a generalised linear model (GLM) with a Gamma distribution and a log link function. To account for potential non-linear relationships, quadratic terms for 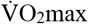were included. The analysis for each fitness estimation method was conducted under two levels of progressive covariate adjustment: 1) adjusted for age, sex, and resting heart rate; and 2) additionally adjusted for height and weight. The overall fit for each method and adjustment level was then compared using the AIC values, where lower values indicate a superior fit. Associations were also visualised by plotting the estimated marginal means of each risk factor across a range of 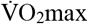 values.

### Statistical software

Statistical analyses were performed with Stata (Version 18; StataCorp, College Station, TX). A p-value of 0.05 or less was considered statistically significant. All code to implement the modelling framework were written in Mata, a C-like matrix programming language that is a component of Stata.

## ABREVIATIONS

ACSM: American College of Sports Medicine
AIC: Akaike information criterion
BMI: Body-mass index
EM: Expectation maximisation
HDL: High-density lipoprotein (HDL) cholesterol
HR: Heart rate
MSE: Mean squared error
MVN: Multivariate normal distribution
PSO: Particle Swarm Optimisation
RMSE: Root mean squared error
SD: Standard deviation
SEM: Structural equation model
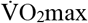: Maximal oxygen consumption

## DATA AVAILABILITY

Biobank validation study, Step test validation study, and Fenland study data are available from the University of Cambridge MRC Epidemiology Unit upon reasonable request: https://www.mrc-epid.cam.ac.uk/research/data-sharing/. Treadmill data from PhysioNet are available at the following repository: https://physionet.org/content/treadmill-exercise-cardioresp/1.0.1/.

## CODE AVAILABILITY

Study code are available upon reasonable request to the corresponding author.

## ACKNOWLEDGEMENTS

We are grateful to all study participants who gave their time and effort. We also thank the functional teams of the MRC Epidemiology Unit at the University of Cambridge (Field Epidemiology, Study Coordination, Data management and IT) for supporting this study.

## AUTHOR CONTRIBUTIONS

T.G. and S.B designed the study. T.G. sourced and pre-processed the data, developed the models, performed the statistical analyses, and produced the figures and tables. T.G. and S.B. wrote the first draft of the manuscript. S.B. is a principal investigator for the Biobank validation study, Step test study, and Fenland study. N.W. is the chief investigator of the Fenland Study. All authors critically reviewed, contributed to the preparation of the manuscript, and approved the final version. All authors vouch for the data, analyses, and interpretations.

## COMPETING INTERESTS

Cambridge Enterprise, the wholly owned subsidiary and technology transfer office of the University of Cambridge, has filed a patent application, GB2411803.6, related to the fitness estimation framework presented in this manuscript with T.G and S.B named as inventors. All authors declare no other competing interests.

## SUPPLEMENTARY TABLES & FIGURES

**Supplementary Table 1.** Agreement statistics for framework-estimated and directly measured 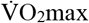 (ml O_2_·min^-1^·kg^-1^) under different data-restriction scenarios. When applying weight and height restrictions, these variables were not used when conditioning the multivariate normal distribution during parameter optimisation. When applying heart rate data restrictions, heart rate greater than 85% age-predicted max heart rate were censored prior to fitting the framework. Age-predicted max heart rate was computed as: 208-0.7·Age(years)

| Restrictions applied when fitting the framework | $\bar{\varepsilon}$ | $\sigma_{\bar{\varepsilon}}$ | RMSE | $r$ | $\rho_c$ | $C_b$ |
| --- | --- | --- | --- | --- | --- | --- |
| No data restrictions applied | 1.5 | 6 | 6.2 | 0.8 | 0.76 | 0.95 |
| Weight and height | 2.1 | 6.6 | 6.9 | 0.76 | 0.69 | 0.91 |
| HR greater than 85% age-predicted max HR | 2 | 6.7 | 7 | 0.76 | 0.73 | 0.97 |
| HR greater than 85% age-predicted max HR, weight, and height | 2.3 | 7.5 | 7.8 | 0.69 | 0.66 | 0.96 |

HR: measured heart rate $\bar{\varepsilon}$ : Mean error, $\sigma_{\bar{\varepsilon}}$ : Standard deviation of the mean error, RMSE: Root mean squared error, $r$ : Pearson's correlation coefficient, $\rho_c$ : Concordance correlation coefficient, $C_b$ : Bias correction factor

**Supplementary Table 2.** Uniform distribution bounds for Particle Swarm Optimisation (PSO) particle initialisation, which defined the range from which each particle’s initial position in the parameter space was sampled.

| Parameter | Lower bound | Upper bound |
| --- | --- | --- |
| $\gamma_1$ | 0.1 | 0.5 |
| $\gamma_2$ | 0.01 | 0.10 |
| $\gamma_3$ | 0.05 | 0.25 |
| $\beta_{Walk}$ | 0.05 | 0.15 |
| $\beta_{1TreadWalk}$ | 0.05 | 0.15 |
| $\beta_{2TreadWalk}$ | 1 | 2 |
| $\beta_{1TreadRun}$ | 0.15 | 0.25 |
| $\beta_{2TreadRun}$ | 0.5 | 1.5 |
| $\beta_{Cycle}$ | 0.1 | 0.2 |
| $\beta_{Step}$ | 0.1 | 0.5 |
| $\dot{V}O_2max$ | 15 | 75 |
| HRmax | 120 | 200 |

**Supplementary Figure 1.**
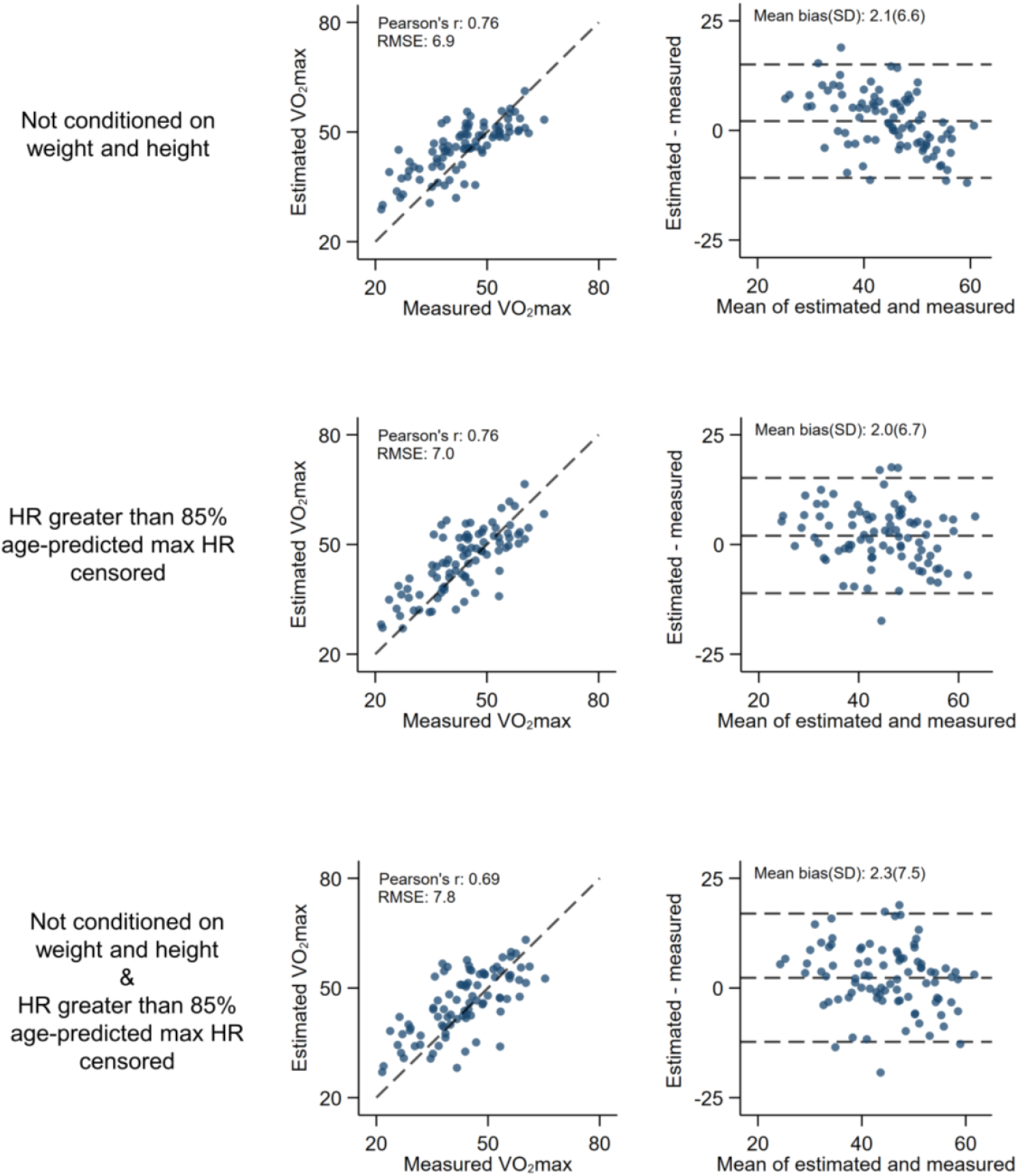
Sensitivity analysis of the framework’s reliance on demographic information. Scatter plots and Bland-Altman plots demonstrate agreement between framework-estimated and directly measured 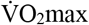 (ml O_2_·min^-1^·kg^-1^) under different data-restriction scenarios. When applying weight and height restrictions, these variables were not used when conditioning the multivariate normal distribution during parameter optimisation. When applying heart rate data restrictions, heart rate greater than 85% age-predicted max heart rate were censored prior to fitting the framework. Age-predicted max heart rate was computed as: 208-0.7·Age(years).

**Supplementary Figure 2.**
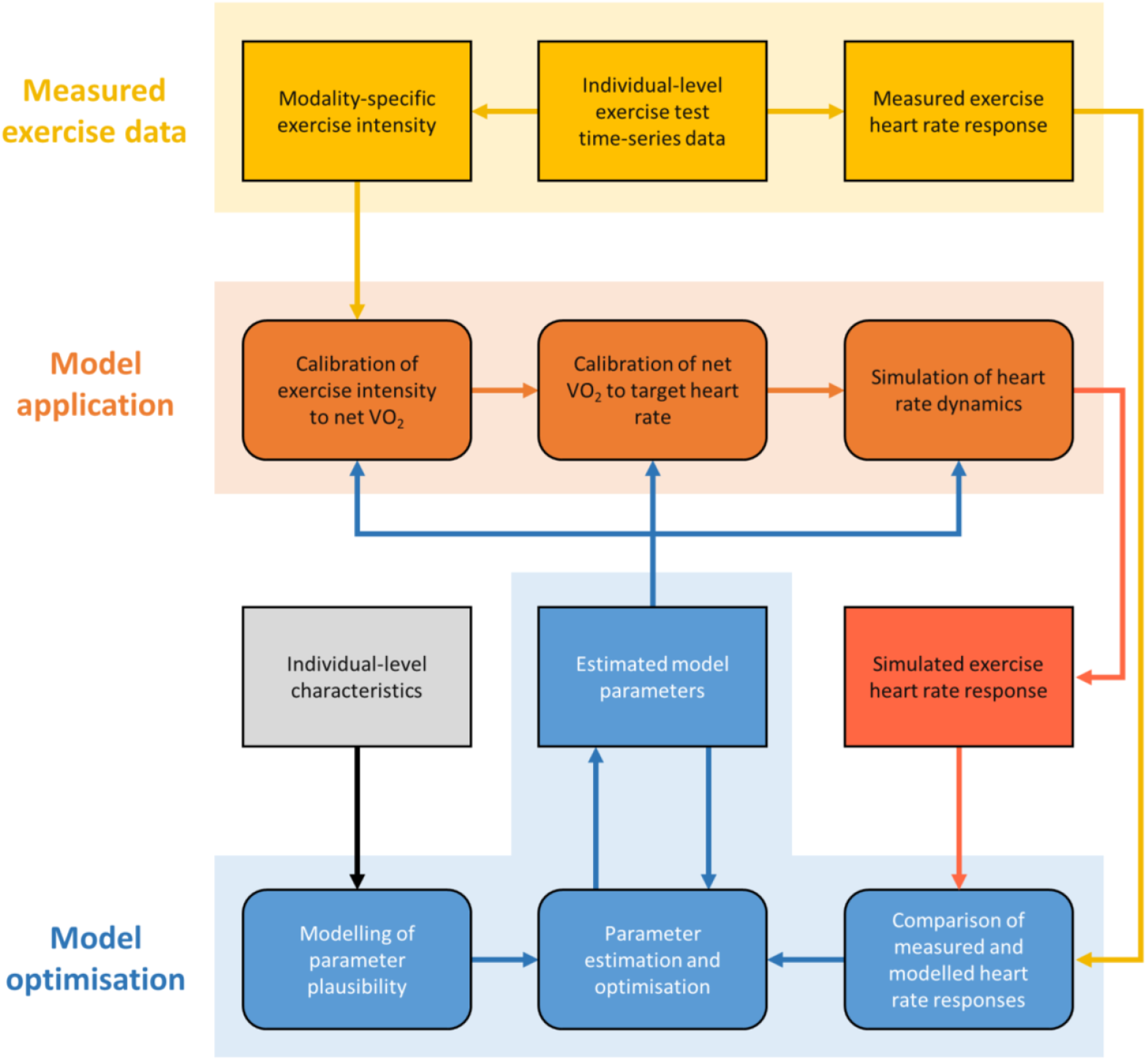
Framework structure for estimating fitness from exercise heart rate response. The framework uses measured exercise data (yellow region), flows to model application (orange region), and then to model optimisation (blue region). Rectangles represent static data elements and parameters. Rounded rectangles represent modelling processes and optimisation steps. Arrows indicate the direction of data flow and the sequence of process interactions. Two-way arrows between “Estimated model parameters” and “Parameter estimation and optimisation” reflect an iterative loop to refine model parameters for both statistical agreement and plausibility. Model parameter outputs include estimated 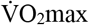, HRmax, and coefficients to calibrate net VO_2_.

**Supplementary Figure 3.**
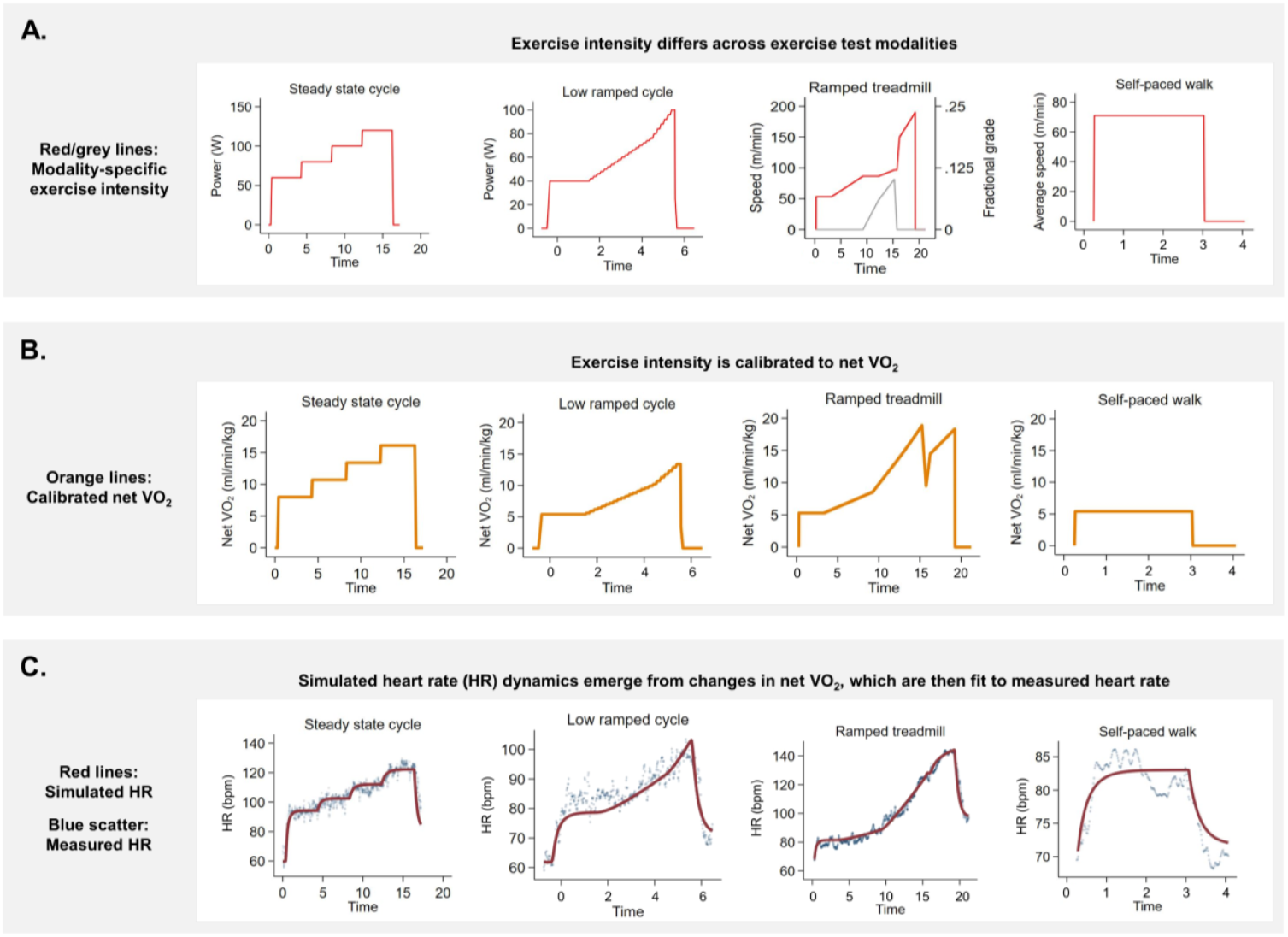
Exemplar plots of data from different exercise tests performed by a single individual demonstrating the framework structure. **A**. Modality specific exercise intensity. **B**. Calibration of exercise intensity to net VO_2_. **C**. Simulation of heart rate dynamics and comparison with measured heart rate. X-axes (Time) represent minute of exercise.

**Supplementary Figure 4.**
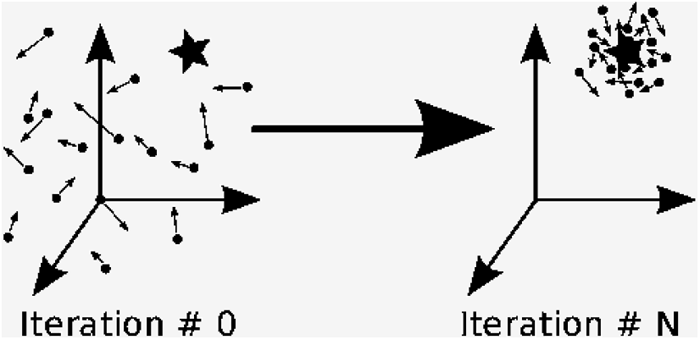
Overview of Particle Swarm Optimization (PSO). The illustration shows the iterative process of PSO. At ‘Iteration # 0’, the particles (dots) are randomly distributed in the parameter space. Each axis corresponds to a parameter being optimised. Arrows projecting from the particles represent their velocities and direction of movement through the parameter space. The star represents the optimal solution. By ‘Iteration # N’, the particles have converged around the optimal solution. **Step 1.** Initialise particles with random positions (parameter values). - pBest (Personal Best): The best position each particle has achieved so far. - gBest (Global Best): The best position achieved by any particle in the swarm. **Step 2.** For each particle: a. Evaluate the objective function at its position. b. Update pBest if the current position has a better objective function value. **Step 3.** Update gBest with the best position found in the swarm based on the objective function. **Step 4.** For each particle: a. Adjust velocity based on its pBest and the swarm’s gBest. b. Update position (parameter values) using this velocity. **Step 5.** If stopping criteria are met (e.g., maximum number of iterations, objective function threshold reached), stop; else, repeat from Step 2.

**Supplementary Figure 5.**
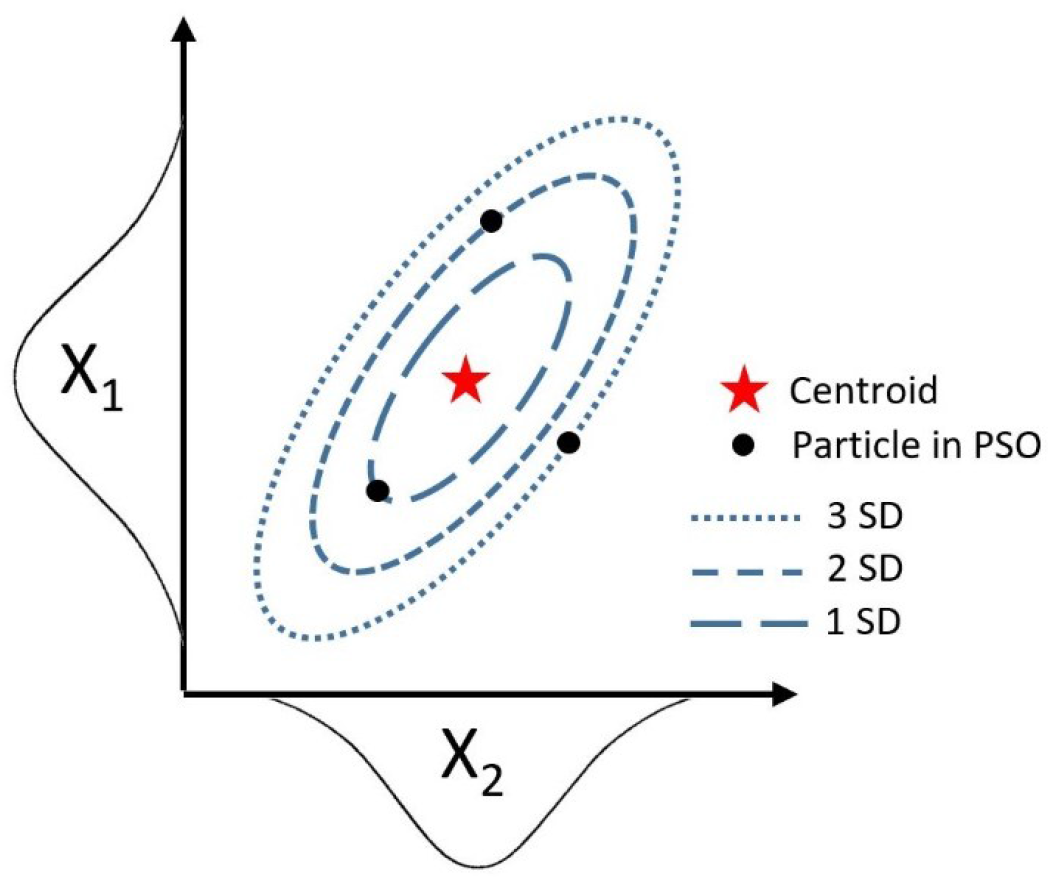
Conceptual figure illustrating the use of Mahalanobis distance in Particle Swarm Optimisation (PSO). Parameters X_1_ and X_2_ are modelled as normal distributions with mean values that intersect at a centroid (red star). Their covariance determines the orientation and elongation of the elliptical dashed contour lines of equal Mahalanobis distance. Particles (black dots) with parameter values closer to the centroid, quantified in standard deviations (SD), have lower Mahalanobis distance. Lower distance values indicate greater physiological plausibility.

## Notes

### Funding Statement

The current work was supported by the Medical Research Council (MC_UU_12015/1, MC_UU_12015/3, MC_UU_00006/1, MC_UU_00006/4), UK Biobank, and the National Institute of Health Research (NIHR) Cambridge Biomedical Research Centre (NIHR203312).

### Author Declarations

Ethical approval was obtained by the University of Cambridge Human Biology Research Ethics Committee (Ref: HBREC/2015.16).

